# The Sanitation Hygiene Infant Nutrition Efficacy (SHINE) Trial: Protocol for School-Age Follow-up

**DOI:** 10.1101/2022.08.17.22278247

**Authors:** Joe D Piper, Clever Mazhanga, Marian Mwapaura, Gloria Mapako, Idah Mapurisa, Tsitsi Mashedze, Eunice Munyama, Maria Kuona, Thombizodwa Mashiri, Kundai Sibanda, Dzidzai Matemavi, Monica Tichagwa, Soneni Nyoni, Asinje Saidi, Manasa Mangwende, Dzivaidzo Chidhanguro, Eddington Mpofu, Joice Tome, Batsirai Mutasa, Bernard Chasekwa, Melanie Smuk, Laura E Smith, Handrea Njovo, Chandiwana Nyachowe, Mary Muchekeza, Kuda Mutasa, Virginia Sauramba, Lisa F Langhaug, Naume V Tavengwa, Melissa J Gladstone, Jonathan C Wells, Elizabeth Allen, Jean H Humphrey, Robert Ntozini, Andrew J Prendergast, the SHINE Follow-up Study Team

## Abstract

**Background:** There is a need for follow-up of early-life stunting intervention trials into childhood to determine their long-term impact. A holistic school-age assessment of health, growth, physical and cognitive function will help to comprehensively characterise the sustained effects of early-life interventions.

**Methods:** The Sanitation Hygiene Infant Nutrition Efficacy (SHINE) trial in rural Zimbabwe assessed the effects of improved infant and young child feeding (IYCF) and/or improved water, sanitation and hygiene (WASH) on stunting and anaemia at 18 months. Among children enrolled to SHINE, 1300 are undergoing follow-up at 7-8 years of age (1000 HIV-unexposed and 300 HIV-exposed children). Children are assessed using the School-Age Health, Activity, Resilience, Anthropometry and Neurocognitive (SAHARAN) toolbox, which provides a holistic measurement of growth, body composition, cognitive and physical function. In parallel, a detailed caregiver questionnaire assesses household demographics, socioeconomic status, adversity, nurturing, caregiver support, food and water insecurity. A monthly morbidity questionnaire is administered by community health workers to evaluate school-age rates of infection and healthcare-seeking. The impact of the SHINE IYCF and WASH interventions, the early-life ‘exposome’, maternal HIV, and contemporary exposures on each school-age outcome will be assessed. We will also undertake an exploratory factor analysis to generate new, simpler metrics for assessment of cognition (COG-SAHARAN), growth (GROW-SAHARAN) and combined growth, cognitive and physical function (SUB-SAHARAN). The SUB-SAHARAN toolbox will be used to conduct annual assessments within the SHINE cohort from ages 8-12 years.

**Ethics and dissemination:** The study has been approved by the Medical Research Council of Zimbabwe (dated 8^th^ February 2021) and registered with Pan-African Clinical Trials Registry (PACTR202201828512110). Primary caregivers provide written informed consent and children written assent. Findings will be disseminated to the community at sensitisation events, through conference presentations, peer-reviewed journals and key stakeholders including the Zimbabwean Ministry of Health and Child Care, and UNICEF.

**Article Summary:**

- This protocol describes the follow-up of school-age children after receiving early-life IYCF and WASH interventions in rural Zimbabwe.
- A comprehensive measurement of the child’s health, growth, body composition, physical and cognitive function will be performed.
- Analysis of the ‘exposome’, including maternal HIV status, will identify the early-life factors that shape school-age physical and cognitive function
- Capitalizing on the randomized trial design will allow the long-term effects of IYCF and WASH on child health outcomes to be estimated.
- Limitations include the re-enrolment of only a subgroup of children from one of the two original study districts; exclusion of HIV-positive children; and the long period between early-life randomized interventions and school-age outcome assessment.

## INTRODUCTION

Children are defined as “stunted” when their height-for-age Z-score (HAZ) is more than 2 standard deviations below the WHO reference standard, but linear growth faltering also affects many children who have not yet fallen below this cutoff^1^. Stunting is associated with increased mortality, poorer school performance, lower adult earnings and long-term chronic disease^2^. Stunting affects 22% (149 million) of children under 5 years^3^, while up to 250 million children are at risk of not reaching their developmental potential^4^. This makes stunting a global health priority, although its causes, longer term impact and response to interventions remain poorly understood.

Given the ongoing global burden of stunting, long-term follow-up studies reflecting contemporary conditions, geography and interventions for stunting are urgently needed to inform robust, cost-effectiveness analyses and advocacy of IYCF and WASH policies and programming (Figure 1). The lack of long-term data evaluating *combined* neurodevelopment, physical fitness and growth has led to a call for further studies that measure a range of outcomes^5 6^. A holistic measurement is vital to understand school-age trajectories across different functional domains and the impact of risk and protective factors and potential interventions (Figure 2). At school-age, it becomes easier to undertake more detailed measures of cognitive development, including school performance, executive function and socio-emotional behaviour.

**Figure 1:**
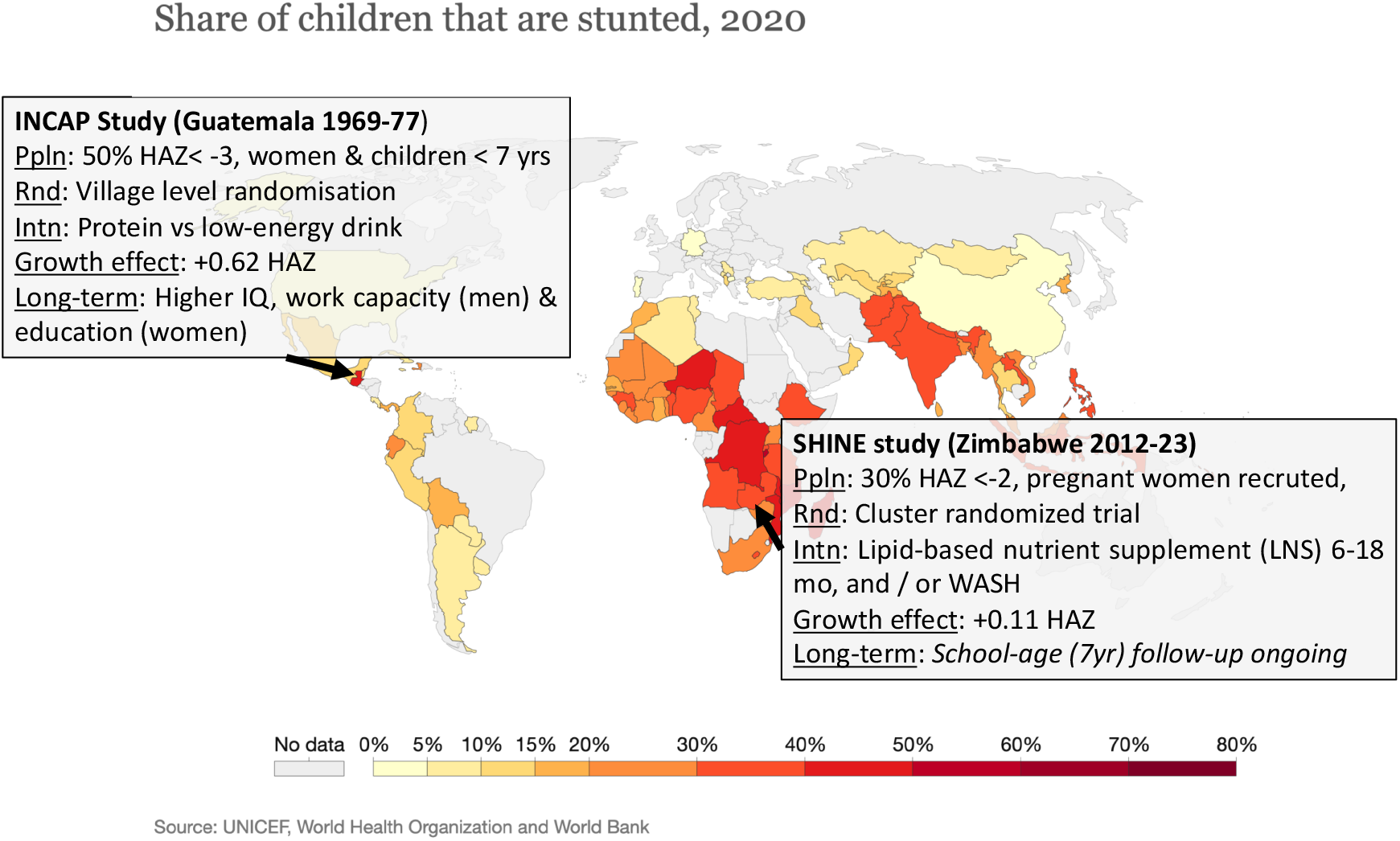
Proportion of children that are stunted (UNICEF, WHO and World Bank, kindly reproduced from ‘Our World in Data’. (Inset details describe the landmark INCAP study and the proposed SHINE study, both of which provide long-term follow-up of a nutrition intervention). Ppln: population, Rnd: randomisation technique, Intn: Intervention. Data from the landmark INCAP study in Guatemala 50 years ago indicate that early-life improvements in nutrition can confer long-term benefits for cognition. Children receiving additional nutrition by age 2 years initially had only modest effects on neurodevelopmental scores; however, in long-term follow-up, those who had received the nutrition intervention had higher IQ scores, greater work capacity and earnings (among men) and greater schooling (among women)^4^. The INCAP study was conducted at a time when global stunting prevalence was much higher (1969-1977): 50% of the study population had HAZ<-3.0, while currently worldwide 22% children have HAZ <-2.0. Furthermore, the impact of the intervention on linear growth was much greater than that seen in trials of complementary feeding interventions over the past 20 years (+0.62 HAZ compared to +0.11 HAZ). Thus, although the Guatemala trial suggests that complementary feeding interventions can have substantial long-term physical and neurodevelopmental benefits, it does not reflect today’s situation in which Africa has the highest stunting prevalence, severe stunting is relatively rare but moderate stunting is 20-40%, and the average impact of interventions on HAZ is only 0.1-0.2.

**Figure 2:**
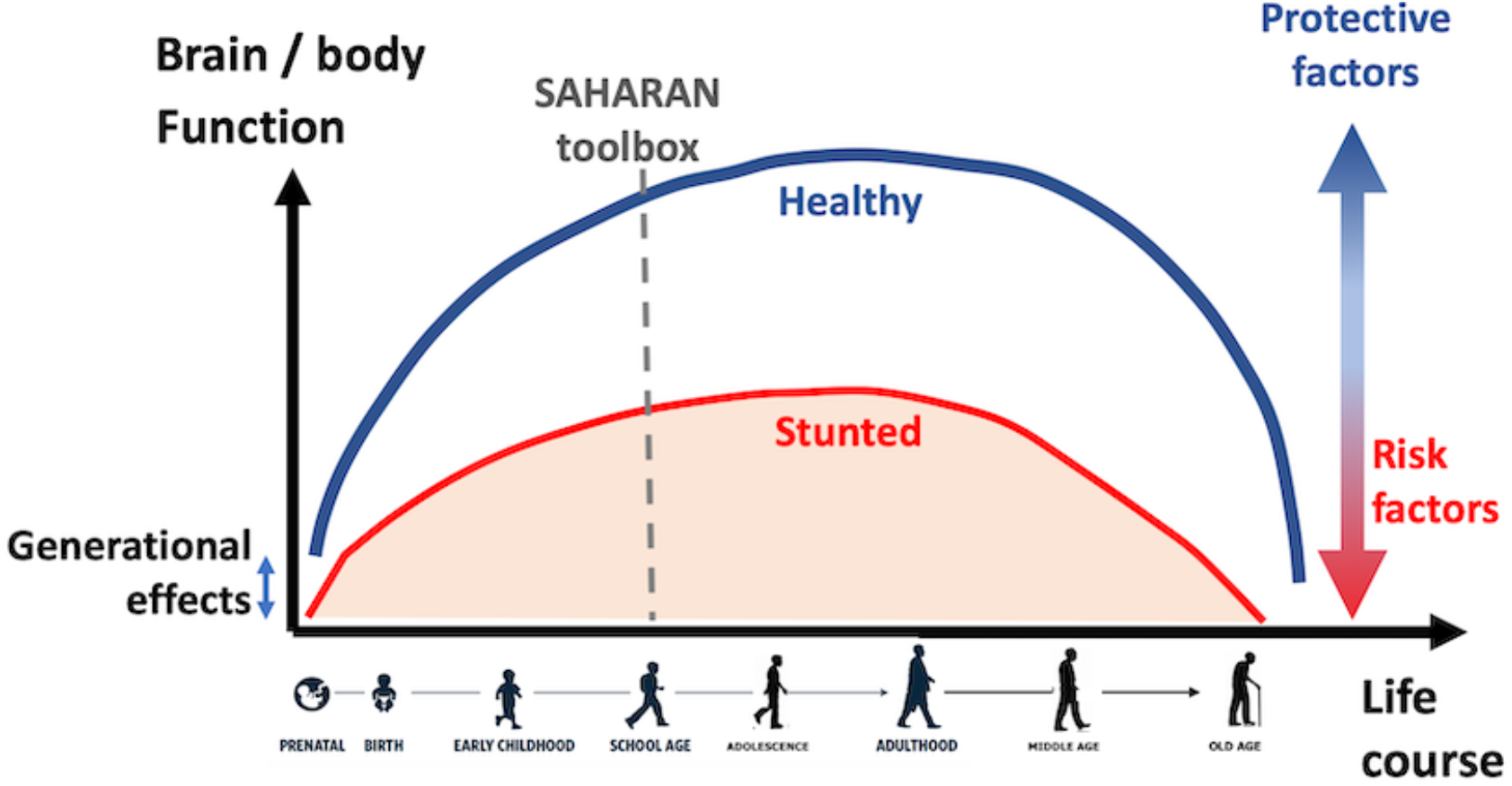
The life course approach to child growth and development, (reproduced from Child Health for all, 6^th^ Edition with permission from Oxford University Press). Healthy (blue) and stunted (red) trajectories are shown, illustrating that lifelong health and function are particularly affected by early-life conditions, as well as risk and protective factors throughout life. This applies to linear growth, physical function and health as well as cognitive function and mental health.

## AIMS AND OBJECTIVES

This study will undertake long-term follow-up of a cohort of children enrolled in the Sanitation Hygiene Infant Nutrition Efficacy (SHINE) trial in rural Zimbabwe. SHINE was a 2×2 factorial cluster-randomized trial across two contiguous districts in rural Zimbabwe between 2012-17, designed to test the independent and combined effects of improved infant and young child feeding (IYCF) and improved water, sanitation and hygiene (WASH) on child height-for-age Z score (HAZ) and haemoglobin at 18 months of age. All outcomes were stratified by maternal HIV status. Detail of the trial design^7^ and primary results^8-11^ are published. In brief, SHINE found that the IYCF intervention modestly improved linear growth, reducing stunting prevalence by 20%, while the WASH intervention had no significant effect. Children who were HIV-exposed and uninfected (CHEU) had a higher risk of stunting and impaired neurodevelopment than children who were HIV-unexposed, but also intriguing evidence of increased responsiveness to early-life interventions^10 11^.

Long-term follow-up of this cohort now enables us to address: i) the effects of the randomised IYCF and WASH interventions on school-age growth, body composition, cognitive and physical function; ii) the impacts of early-life stunting and a range of other exposures (the ‘exposome’) on school-age growth, body composition, cognitive and physical function; and iii) the differences in school-age growth, body composition, cognitive and physical function between HIV-exposed and HIV-unexposed children. Therefore the objectives are summarised as:

1. Evaluate the impact of the IYCF intervention on school-age growth, health, physical and cognitive function (both for HIV-exposed and unexposed children)
2. Evaluate the impact of the WASH intervention on school-age growth, health, physical and cognitive function (both for HIV-exposed and unexposed children)
3. Evaluate the relationship between the exposome during the first 1000 days and school-age growth, health, physical and cognitive function, including:
  a. Early-life length-for-age Z-score, and categorical definitions of stunting (LAZ<-2) by 1 month of age (early stunting) and by 18 months of age (late stunting).
  b. Environmental factors, including socioeconomic status, household composition, demographics, maternal education, household adversities, food and water insecurity.
  c. Pregnancy exposures including maternal caregiving capabilities, nutritional status and depression.
  d. HIV exposure in pregnancy.
4. Evaluate the relationship between current environmental, schooling, nurturing and care-giving practices and school-age growth, health, physical and cognitive function.
5. Develop and deploy three novel, holistic and succinct *outcome* metrics to assess school-age growth, health, physical and cognitive function.

## METHODS

SHINE enrolled pregnant women from 210 clusters^7 8^, each representing the catchment area of 1-4 community health workers (CHW), and covering a geographic area of 8232 km^2^. For this long-term follow-up study, up to 1300 children born to these women from 105 clusters in Shurugwi district will be enrolled. Children who originally participated in SHINE and who are aged between 7-8 years during the period of enrolment, and are HIV-unexposed (CHU) will be randomly selected by computer from clusters by the data team, for a total of 1000 children not lost to follow-up. All HIV-exposed children will be invited to enrol, aiming for up to 300 CHEU in total; any child testing HIV-positive will be excluded from analysis. Children no longer resident in Shurugwi, with unknown maternal pregnancy HIV status, or outside the age window will be ineligible. There are no new interventions in the long-term follow-up study.

### Community sensitisation and pre-screening

At the start of the follow-up study, sensitisation of the local leadership and community members was undertaken by the study team, together with the screening of a community-made film about the SHINE trial which explored their previous involvement and experiences of research. CHWs performed role-plays to explain the purpose, consent procedures and practicalities of the visit to participating families and the community. Once the family has received information and expressed an interest in joining, a mutually convenient date is booked for an assessment.

### Screening, consent, assent and assessments

Assessments are conducted by data collectors (DCs), who are primary care nurses extensively trained in study procedures. On the day of the assessment, the CHW introduces a pair of DCs (DC1 and DC2) to the household. Screening is undertaken to ensure the original SHINE child and primary caregiver are present; once both DCs verify that this is the correct child, 1-2 tents are pitched in or close to the household (Figure 3). Households are usually single-family dwellings surrounded by subsistence farmland and the mean distance between houses in SHINE was 82.6m^8^, ensuring the tents are sufficiently isolated for confidentiality during informed consent and data collection. Written informed consent is obtained from the primary caregiver, and assent from the child, following a demonstration of the tools used to conduct the measurements. The primary caregiver is usually the mother, but it is the child born within the SHINE trial who is the focus of this follow-up study. Hence if the mother is not available (for example the mother has died, divorced or moved away) the primary caregiver is defined as the adult most responsible for the child’s care, and completes the caregiver questionnaire.

**Figure 3.** [Photo]: SHINE school-age follow-up: a) Measurement tent erected in household yard b) Cognitive measurement showing the school achievement test (SAT) c) Body composition measurement showing subscapular skinfold d) Physical function measurement showing shuttle run test.

DC1 performs the caregiver questionnaires in a pre-specified order, while DC2 conducts the cognitive, physical and anthropometry assessments in a separate tent to minimize distractions, after building rapport with the child. The child is fed before the start of the tests (either by the family or the DC’s who provide additional snacks). The child is then given further regular breaks and snacks to reduce tiredness. All tests have standardized explanations and demonstrations to ensure appropriate understanding, and are translated into Shona or Ndebele – the local languages spoken in the district. The DCs remain blinded to the original trial allocation and the maternal HIV status throughout the visit. At the end of the visit, DC1 opens a sealed envelope to check the maternal HIV status in the original trial, which then determines whether questions are asked on HIV treatment or if HIV testing is appropriate. At the end of the visit, a small gift is given to the child and caregiver to thank them for their time.

### HIV testing and referrals

All primary caregivers are offered HIV testing at the end of the visit, unless they are already known to be living with HIV or had a documented negative result in the previous 3 months. If the mother is HIV-negative, the child is not tested. If the mother is HIV-positive, declines testing or is not available, the child is offered HIV testing with age-appropriate assent using role-plays. The Determine HIV-1/2 rapid test (Abbott) is used for initial testing; positive results are repeated using the HIV 1/2 Stat-Pak rapid test (Chembio). The option of dried blood spot testing is also available for participants who prefer not to be tested in real-time in the homestead. Referrals are made to local clinics for mothers or children testing HIV-positive.

## OUTCOME MEASURES

All assessment tools have been integrated into a single assessment lasting 4-5 hours, using the School Age Health, Activity, Resilience, Anthropometry and Neurocognitive (SAHARAN) toolbox, which was developed, refined and piloted by our team^12^. Briefly, SAHARAN includes a caregiver questionnaire, child questionnaire and direct tests undertaken with the child, focused on three domains: cognitive function, growth and body composition, and physical function. The primary outcome of the long-term follow-up study is cognitive function, assessed by the mental processing index (MPI) - the total score from 8 subtests of the Kaufman Assessment Battery for Children 2^nd^ edition (KABC-II). The subtests measure four domains of cognitive processing across learning, planning, simultaneous and sequential memory. All other outcomes are secondary outcomes and shown in Table 1.

**Table 1:** Cognitive outcomes to be measured, including the primary outcome (MPI) and secondary outcomes.

|  | MARKER | MEASURE | OUTCOMES | RATIONALE |
| --- | --- | --- | --- | --- |
| Cognitive Function (120 mins including KABC-II) | Kaufman Assessment Battery for Children | Cognitive processing | Primary outcome: Mental processing index (MPI) | Overall measure of cognitive function |
|  |  |  | Secondary outcomes: KABC-II domain scores, individual subtest scores | Short & long-term memory, planning, problem-solving, sequential memory |
|  | School Achievement Test | Academic | Total score, Subtest scores (*numeracy, reading, writing*) | Literacy & numeracy |
|  | Fine motor | Shortest time to complete finger tapping sequence | Time for dominant hand, non-dominant hand, and average between both hands, | Fine motor |
|  | Plus-EF Tablet test | Executive Function | Overall score, individual subtest scores, reaction time | Executive function |
|  | Child socio-emotional questionnaire | Home support | Total score, sub-score removing food insecurity question | Child's own perspective on home support |
|  | Washington Group Child function module (asked in caregiver questionnaire) | Disability screening, including vision and hearing | Overall score, disability, learning, mental health subscale | Child functional abilities |
|  | Strength and Difficulties Questionnaire (SDQ, asked in caregiver questionnaire) | Socioemotional function | SDQ total score and subtest scores | Behaviour |

**Table 2:** Growth and body composition outcomes. MUAC: (Mid upper-arm circumference)

|  | MARKER | MEASURE | OUTCOMES (secondary) | RATIONALE |
| --- | --- | --- | --- | --- |
| Body composition (20 mins) | Bio-impedance analysis (BIA) | Impedance of tissues | Lean mass index, Phase angle, Impedance index, Reactance, Resistance | Quality of growth, metabolic health |
|  | Knee-heel length | Tibial growth | Median Knee-heel length | Prioritization of growth |
|  | Triceps, scapular, supra-iliac, calf skinfolds | Subcutaneous fat | Sum of skinfolds, Individual skinfolds, Peripheral: central skinfolds, | Subcutaneous fat: peripheral vs. central, metabolic health |
| Anthropometry (15 mins) | Height, weight | Growth | HAZ, WAZ, BMI | Growth, nutritional status, Metabolic health |
|  | Head circumference | Brain volume | Head circumference | Prioritization of growth |
|  | Waist and hip circumference | Abdominal size | Waist circumference, hip circumference | Nutritional status, metabolic health |
|  | Calf circumference, MUAC | Peripheral fat & muscle | Calf circumference, MUAC | Quality of growth |

### Support and supervision

Ongoing support and monitoring are provided by a senior research nurse and a doctor, with 6-monthly standardisation exercises. Additional training is provided for any DCs with coefficients of reliability (‘R’) below a set threshold in anthropometry or cognitive measurement, derived from the intra- and inter-observer technical error of measurement for continuous measurements (e.g. anthropometry)^13^ and intraclass correlation coefficient (ICC) for discrete measurements (e.g. cognitive test scores).

### Measurement of illness and healthcare-seeking behaviour

The incidence of common infections is measured by prospective surveillance. The CHW visits families 4-weekly to conduct a 7-day illness recall questionnaire to capture episodes of fever, cough, diarrhoea, skin, ear or mouth infections or other illnesses, and associated health-seeking behaviours.

## SAMPLE SIZE

The primary outcome is the total KABC-II from all subtests, called the Mental processing index (MPI). The raw subtest scores are adjusted by age into 4-month blocks. 1000 children (500 IYCF vs 500 non-IYCF) will be assessed, providing 86% power to detect a 0.2 standard deviation difference in MPI between IYCF and non-IYCF arms with alpha 0.05, assuming intra-cluster correlation of 0.05 and sampling from 100 clusters. This will allow the exploration of the difference in IQ scores observed at 3-7 years of age among children followed-up in the INCAP study, and is also the approximate magnitude of socio-emotional difference recently shown with a similar small-quantity lipid-based nutrient supplement trial^14^. Since data collection is undertaken directly after consent and enrolment, no adjustment for lost to follow-up is required. The SHINE 2×2 factorial design also enables 500 WASH versus 500 non-WASH children to be assessed as a secondary outcome. For CHEU, the target sample size is 300, which will allow exploration of the association between the treatment arms (IYCF vs non-IYCF, and WASH vs non-WASH) for secondary outcomes.

## DATA MANAGEMENT

DCs electronically collect the SAHARAN questionnaire and observation data onto password-protected Android tablets (Samsung Galaxy Tab A) using the Open Data Kit (ODK) platform (https://opendatakit.org/). The ODK forms are programmed with skip patterns, expected data ranges and free text sections to report any issues with data collection. Back-up paper forms are also provided in case of tablet failure. New data are manually checked by the Project Lead and Field Data Officer before being uploaded onto an ODK Aggregate Server, and stored on an SQL Server database at the end of each working day. All data are collected using a unique participant identifier (PID) to maintain confidentiality.

## STATISTICAL ANALYSIS

A Statistical Analysis Plan (SAP) designed to give transparency to possible influential statistics decisions for the study will be finalised prior to unblinding and initiation of analyses. Reporting of results will follow the guidelines established in the extended CONSORT guidance for cluster-randomized trials^15^. The analyses are described below according to the objectives.

### Effect of IYCF and WASH on outcomes (Objectives 1 & 2)

We will capitalize on the 2×2 factorial trial design to evaluate the IYCF and WASH interventions as two trials run in the same population, stratified by maternal HIV status. For the analysis of IYCF as the primary outcome, we will therefore combine the two IYCF-containing trial arms (IYCF alone, and IYCF+WASH) and compare them to the two non-IYCF arms (WASH alone, and standard-of-care). For the analysis of WASH as a secondary outcome, we will combine the two WASH-containing trial arms (WASH alone, and IYCF+WASH) and compare them to the two non-WASH arms (IYCF alone, and standard-of-care). The effect of IYCF and WASH on each outcome (as defined in Tables 1 to 3) will be evaluated using generalized estimating equations (GEE) with an exchangeable working correlation structure to account for within-cluster correlation, assuming no interactions between randomised interventions. All analyses will account for the nature of the distribution of the outcome and results will be presented as appropriate effects sizes (mean difference between groups and risk ratios) with a measure of precision (95% confidence intervals). Analyses will be by intention-to-treat at the child level, according to the mother’s assigned study arm based on her residence at the time of her enrollment into SHINE, regardless of subsequent moving or adherence to the interventions. The hypothesis test will be 2-sided and a significance level of 5% will be used.

The primary analysis will be unadjusted. A secondary adjusted analysis will account for original trial stratification factors and other possible confounders as defined in the SAP. Since we are performing a large number of tests, we will base our conclusions on associations observed in outcome groupings, and not on p-values. The generalisability of the sample will be assessed in comparison to the original SHINE cohort. If evidence of a large difference is seen, then weights will be applied as a sensitivity analysis to explore inference effect.

#### Evaluating early-life growth and exposures on school-age function (Objectives 3a, b, c)

As secondary outcomes, we will explore the effects of early stunting (by 1 month of age) and late stunting (by 18 months of age) on each outcome at 7 years of age, and the effect of LAZ as a continuous variable at 1 and 18 months. For the early-life exposome, we will follow recommended approaches for large datasets, and use principal components analyses to first reduce the number of exposure variables into a smaller set of functional biological domains that best represent the most informative combinations of exposures. We will then use appropriate techniques to quantify the effects of pregnancy and baseline exposures on school-age outcomes^16^, exploiting the rich SHINE dataset to control for confounding by pregnancy-related factors (e.g. wealth, maternal education, haemoglobin, maternal capabilities), the trial interventions, and the time-varying nature of exposures experienced between birth and 18 months of age.

#### CHEU and CHU comparison (Objective 3d)

Baseline characteristics between CHEU and CHU groups will be compared using multinomial and ordinal regression models and Somers’ D for medians, while handling within-cluster correlation with robust variance estimation. We will use GEE models to compare each functional outcome, assessed using the SAHARAN Toolbox, between CHEU and CHUU groups. In addition, we will compare CHEU and CHU using the reduced school-age outcomes and early-life exposures outlined above. A secondary adjusted analysis will be performed including possible confounders as defined in the SAP. Poisson regression will be used to compare the cumulative incidence of illness episodes, clinic visits, and hospitalizations between groups during prospective surveillance measured by the CHW illness questionnaire.

#### Comparison between aspects of school-age growth and function and contemporary conditions (Objective 4)

Internal consistency of outcomes across cognitive domains will be compared, including the primary outcome (KABC-II total score) and other cognitive measurements. Similarly, consistency of physical function measurements and their associations with growth will also be measured, as previously described^12^. Univariable analysis will explore associations between school-age growth, cognitive and physical function (Tables 1 to 3) and current environmental, schooling, nurturing and caregiving exposures (Table 4). We will also undertake an exploratory factor analysis to identify the most informative combinations of measures from the SAHARAN Toolbox. To guide biological interpretation and identify relevant functional domains from the results of the data-reduction step, we will explore factor loadings using bi-plots. We will also apply hierarchical clustering of principal component scores to group children according to their school-age outcomes. We will explore univariable distributions (central tendencies, variance, prevalence) of school-age outcome variables within each identified cluster of children to gain further insights into trade-offs and prioritisation in growth and function^17^.

**Table 3:** Physical function outcomes.

|  | MARKER | MEASURE | OUTCOMES (secondary) | RATIONALE |
| --- | --- | --- | --- | --- |
| Physical Function (30 mins) | Grip strength (a) | Lean muscle both hand | Highest grip strength, Standardised grip strength (a) Dominant and non-dominant hand strength, | Lean muscle: hand |
|  | Broad jump (b) | Truncal muscles | Maximum distance, standardised distance (b) | Lean muscle: leg |
|  | 20m Beep test (c) (composite score = standardised a+b+c) | Physical Fitness, | Shuttle run test level Standardised shuttle run test level: (c) **Composite standardised score = a+b+c** | Stamina, Overall composite score |
|  | Haemoglobin | Anaemia | Hb | Physical fitness |
|  | Blood pressure (BP) | Fitness | Resting Systolic & diastolic BP, Pulse pressure, systolic & diastolic BP & pulse pressure 1 minute after exercise; Post-exercise difference between 1<sup>st</sup> and 5<sup>th</sup> BP systolic & diastolic measurement | Cardiovascular fitness |

**Table 4:**
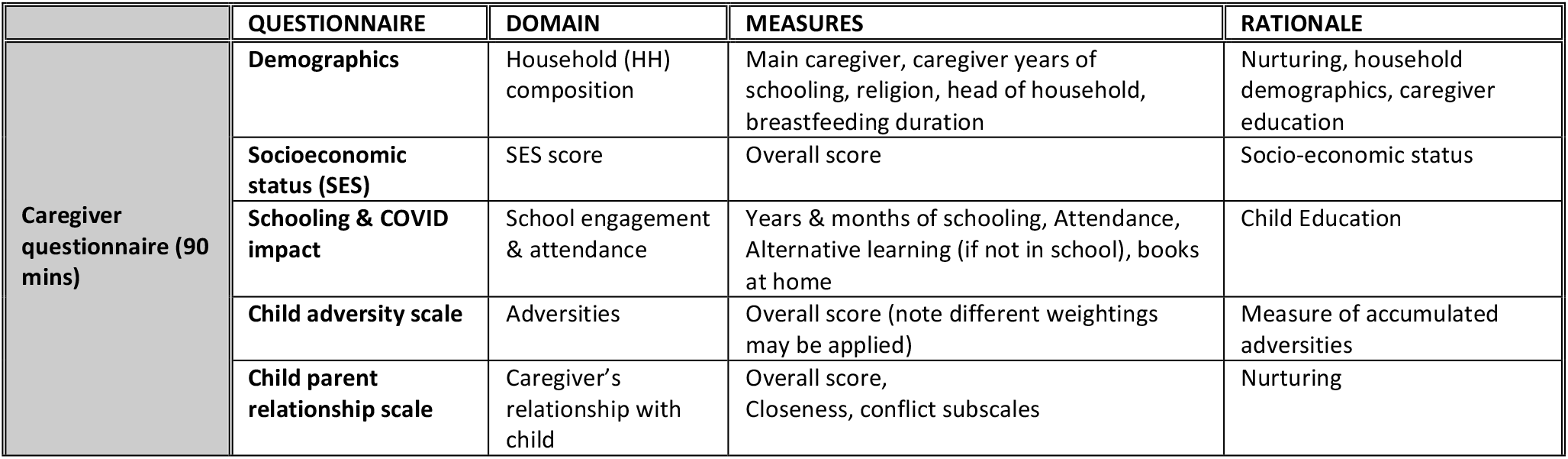
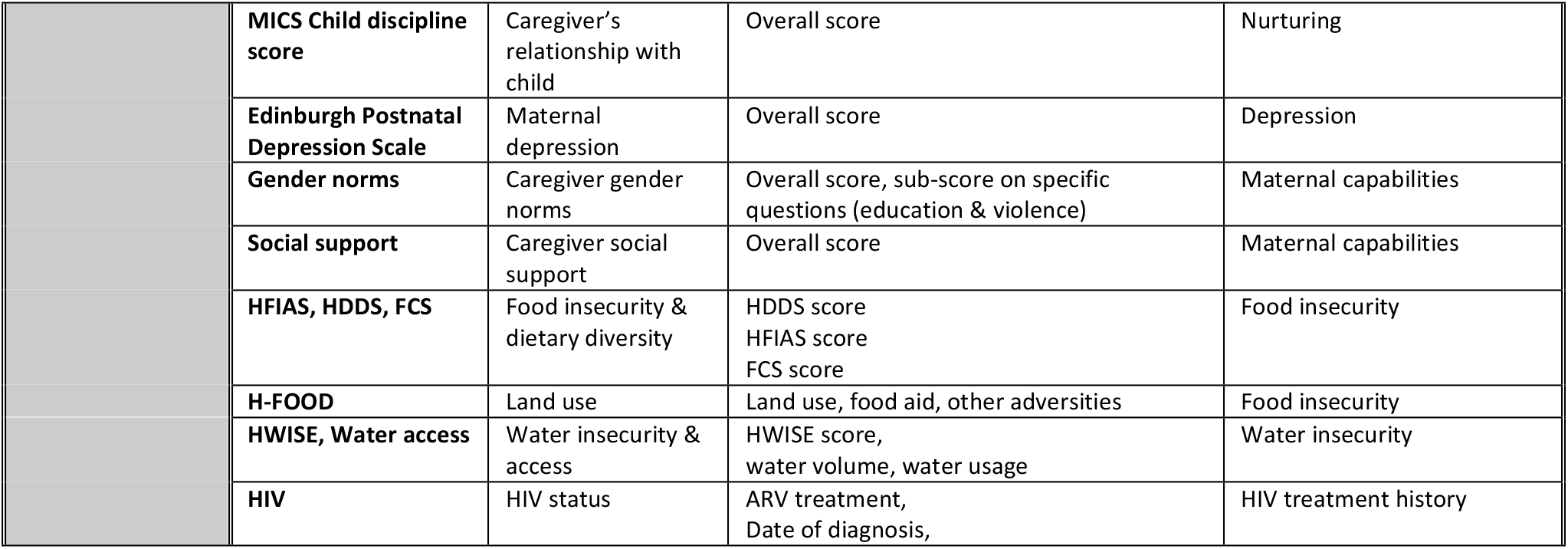
Caregiver questionnaire contemporary exposures. MICS: Multi-indicator cluster survey from UNICEF, HFIAS: Household Food Insecurity Assessment Scale, HDDS: Household Dietary Diversity Score, FCS: Food Consumption Score, HWISE: Household Water Insecurity Experiences Scale Note: WG & SDQ sections are included in cognitive outcomes.

### Development of novel outcome metrics (Objective 5)

Novel outcome metrics will be developed based on SAHARAN toolbox measurements conducted in 250 HUU children in the standard-of-care arm. We will use these data to undertake a factor analysis of the primary and secondary cognitive SAHARAN variables to select the most informative sparse combination of tests for development of a cognition metric (COG-SAHARAN). We will undertake reliability and agreement analyses to compare the performance of the shortened, open-access tools with the KABC-II tool, which is a gold-standard assessment, but is costly, time-consuming, complex and proprietary. A similar factor and reliability analysis will inform the generation of the growth metric (GROW-SAHARAN) from all growth and physical function outcomes. GROW-SAHARAN will measure the child’s growth, body composition and physical function to holistically assess school-age children’s nutritional status. Further analyses could tailor the GROW-SAHARAN to focus on key indicators within specific areas such as physical function, body composition or chronic disease risk. This is a sufficient sample size to derive the COG-SAHARAN and GROW-SAHARAN metrics, based on previous published metrics^18 19^. As outlined above, we will employ factor analysis as a data reduction step to i) identify the underlying constructs that we are measuring, ii) evaluate which of the multiple sub-tests are driving the variability measured, and iii) determine whether all tests are required.

Standardisation exercises with the same children will allow us to test the reliability of the proposed measures using intra-cluster correlation coefficients and to measure Cronbach’s alpha. Finally, the relationship between these novel metrics will be investigated within a recognised conceptual framework that evaluates child function^20^ (Figure 4). A combined child metric (“SUB-SAHARAN”) will also be developed after derivation of COG- and GROW-SAHARAN, by exploring the relationships between the two metrics. All metrics will then be operationalised in the 1000 CHU randomized to early-life IYCF or WASH interventions, and the performance of each metric compared to the more detailed analysis outlined in previous sections.

**Figure 4:**
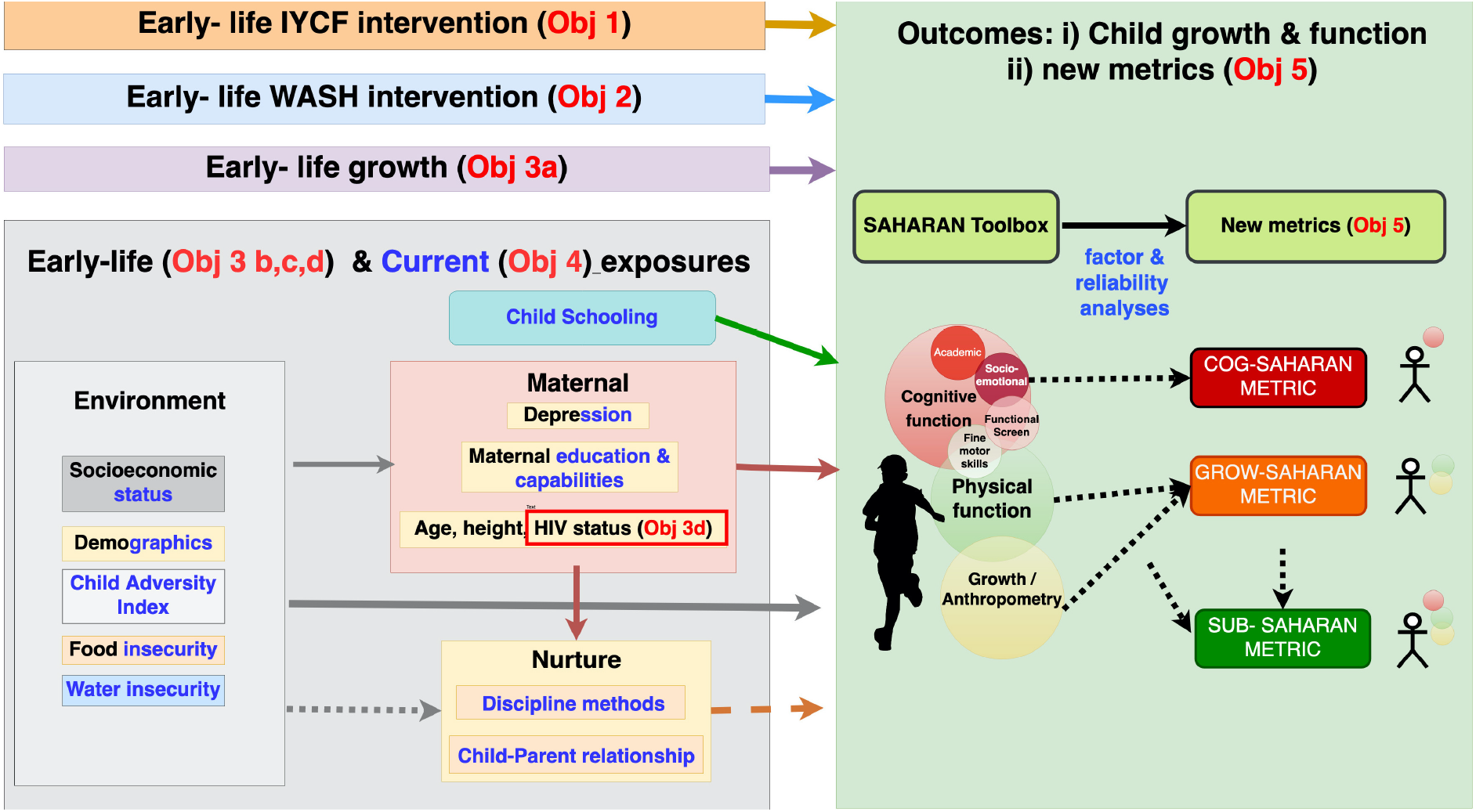
Conceptual framework of objectives with exposure and outcome variables. Exposures are split into environmental, schooling, maternal and nurturing domains. Early-life exposures are described in black and contemporary exposures in blue text. Those exposures that were measured in both early-life and contemporary are written in black and blue text. Outcomes are based on the SAHARAN toolbox to provide school-age child growth, health and function. These outcomes will also be analysed in the standard of care arm to provide new cognitive (COG-SAHARAN), Growth (GRO-SAHARAN) and overall (SUB-SAHARAN) metrics.

## DISSEMINATION

Results of the follow-up study will be disseminated through community gatherings, building on recent screenings of a locally made film about the SHINE trial. We will also leverage the district Community Engagement Advisory Board, which comprises key stakeholders from the community. Findings will be disseminated to the academic community through open-access peer-reviewed journals and conference presentations. In addition, a short animation video describing the school-age assessment metrics will be created and published online, as well as workshops with key local and national partners including Ministry of Health and Child Care, UNICEF, Food and Agriculture Organisation, World Food Program, Ag2Nut and the Agriculture, Nutrition and Health (ANH) academy.

## ETHICS

All research procedures are conducted in accordance with the Declaration of Helsinki and according to International Conference on Harmonisation GCP guidelines. The Medical Research Council of Zimbabwe (MRCZ) approved the study protocol. Adverse events and serious adverse events that are related to any trial procedures during the follow-up visits are reported to MRCZ. Internal monitoring is conducted, and findings requiring corrective action are categorised as critical, major, or minor, with appropriate timelines for resolution. This follow-up study is registered with the Pan-African Clinical Trials Registry (PACTR202201828512110).

## DISCUSSION

### The ‘missing middle’ of school-age child health and function

The SHINE follow-up study aims to characterize school-age health and function and provide important insights into the ‘missing middle’ of childhood. It is known that between 5-14 years, mortality is concentrated in sub-Saharan Africa^21^, but measures of child growth and development have frequently been overlooked^22^. Nevertheless, this life-stage remains crucial for growth to consolidate positive gains from early childhood. Height and BMI monitoring shows that children can experience negative trajectories of increasing BMI in response to adverse environmental conditions^23^, increasing future non-communicable disease risk. Examining growth and development together is urgently needed to understand the longer-term impact of early-life exposures and interventions in relation to school-age growth and function^6^. Few studies have confirmed whether early growth gains translate into long-term improvements in cognitive and physical function and, conversely, whether interventions showing little or no obvious effect at early ages may confer meaningful health and functional benefits in later childhood. It is also important to understand the contemporary risk and protective factors that undermine or accelerate progress, respectively (Figure 2). School-age presents an opportunity to mitigate early disadvantages and consolidate early gains, thus providing sustainable benefits in development and longer-term health.

### Strengths and Limitations of this study

Follow-up of the SHINE cohort at 7 years of age will determine whether nutrition interventions in the first 1000 days that modestly increase linear growth can restore long-term function, and whether WASH interventions in the first 1000 days have any impact on school-age outcomes, despite no effects on early-life growth. This will be achieved using the SAHARAN toolbox to measure growth, neurodevelopmental and physical health outcomes of school-age children, which has previously been extensively piloted. The study efficiently capitalizes on the factorial trial design to evaluate both interventions in the same children. The feasibility of a similar detailed battery in children has previously been demonstrated before in a cohort study of survivors of severe acute malnutrition^24^. Limitations include the selection of only one of the two districts of the SHINE trial, as well as the inability to follow-up children who have moved out of Shurugwi district. The selection of HIV-unexposed children is randomised to ensure balance between the SHINE intervention arms. However, we will try to follow all HIV-exposed children, which may lead to an imbalance in treatment arms for CHEU. HIV-positive children are excluded from the follow-up because there will be too few children to provide meaningful inferences. Finally, all families received a latrine following the 18-month primary endpoint visit, so the focus on interventions is restricted to early-life, whilst recognising the long period between early-life randomized interventions and school-age outcome assessment.

### Effects of HIV exposure on school-age outcomes

There remains an urgent need to clarify whether the health gap between CHEU and CHU in sub-Saharan Africa persists at school age, and to identify the biological, social and environmental exposures in early life that underpin these differences. There is a lack of long-term data: one study in Zambia found that the gap between CHEU and CHU growth outcomes had widened by 7.5 years of age^25^, while a study of neurocognitive outcomes across five African countries^26^ found no differences between CHEU and CHU groups, but did not compare language, which may be most predictive of future function^26^. Other cohorts have reported poorer mathematic abilities and reduced IQ among CHEU in south-east Asia^27^, but studies have focused on a limited range of outcomes. Therefore, applying the SAHARAN toolbox to provide concurrent assessments of growth, health, physical capacity and cognition will allow a holistic insight into the long-term effects of HIV, ART and other pregnancy exposures on health and human capital.

### Derivation and application of metrics

Rural Zimbabwe represents a setting of subsistence farming, food and water insecurity, gender inequity, climate change and political/economic fragility. Measuring these environmental exposures, as well as other household and individual factors, enables their effects on school-age growth and function to be investigated. The SAHARAN toolbox takes approximately 4 hours to complete in the field, which is not feasible for widespread applicability. However, using these data to generate readily accessible and applicable shorter metrics will provide future tools that rapidly assess either school-age cognitive function (COG-SAHARAN), physical function and growth (GROW-SAHARAN), or both (SUB-SAHARAN). These metrics will be derived from indicators and tests previously used in other LMIC and hence should be widely applicable. We will provide a toolkit for operationalisation of the metrics in other contexts, allowing other research groups and programmes to adapt the tools prior to implementation.

## Conclusion

Follow-up of the SHINE trial provides a unique opportunity to comprehensively characterise the long-term response to early-life nutrition and WASH interventions, for both HIV-exposed and HIV-unexposed children. In this context, use of the SAHARAN toolbox provides a holistic outcome measurement which is crucial to understand school-age trajectories across different functional domains and the long-term impact of exposures including interventions, risk and protective factors. Our study will inform future policy and programming approaches to stunting, since demonstration of long-term functional benefits would motivate scale-up of IYCF in areas of high stunting or HIV-exposure. Conversely, a lack of school-age benefits suggests that alternative approaches to stunting, poor neurodevelopment and their subsequent mitigating strategies are urgently needed. The metrics themselves also represent paradigm shifts in evaluation of nutrition-sensitive interventions in a broader context; they will provide assessment tools for school-age growth and non-communicable disease risk, with functional measures of physical health and human capital. It is only by generating both the measurement tools and long-term data that evidence can be translated into action.

## Data Availability

All data produced in the present study are available upon reasonable request to the authors

https://osf.io/w93hy/

## Data statement

The study protocol and all data collection tools are available on Open Science Framework https://osf.io/w93hy/

Statistical code, and dataset will be available from the data repository, https://clinepidb.org/ce/app

## Author Contributions

Joe D Piper, Robert Ntozini, Jean Humphrey and Andrew J Prendergast conceived the study, designed, wrote and registered the protocol, gained ethical approval, coordinated data collection, undertook data analysis, interpretation, write-up and dissemination. Clever Mazhanga coordinated pretesting, translations, sensitisation, training, data collection and provided feedback and critical input to the study design. Marian Mwapaura coordinated data collection and performed visit planning and data management. Gloria Mapako, Idah Mapurisa, Tsitsi Mashedze, Eunice Munyama, Maria Kuona, Tsitsi Mashedze, Thomizodwa Mashiri, Kundai Sibanda, Monica Tichagwa, Nyoni Soneni, Asinje Saidi andManasa Mangwende performed pretesting, sensitisation, data collection and provided feedback and critical input to the measurement tools. Dzivaidzo Chidhanguro, Naume V Tavengwa and Lisa F Langhaug coordinated study planning, sensitisation, field operations, data collection and provided feedback on the study design. Eddington Mpofu, Joice Tome and Batsirai Mutasa contributed data collection tools and data management. Handrea Njovo and Chandiwana Nyachowe contributed to the study design and provided support from the Zimbabwean Ministry of Health and Child Care. Melissa J Gladstone contributed study design, choice and application of cognition tools, data interpretation and write-up. Jonathan C Wells contributed study design, choice and application of body composition tools, data analysis, interpretation and write-up. Melanie Smuk, Bernard Chasekwa and Robert Ntozini provided statistical planning, analysis, randomisation and support. All authors read the manuscript, contributed revisions as required and approved the final manuscript.

## Acknowledgements

We thank Zvitambo staff, the Ministry of Health and Child Care, and the community leadership in Shurugwi for their help with this study.

## REFERENCES

1. Perumal N, Bassani DG, Roth DE. Use and Misuse of Stunting as a Measure of Child Health. The Journal of nutrition 2018;148(3):311–15. doi: 10.1093/jn/nxx064

2. Martorell R. Improved Nutrition in the First 1000 Days and Adult Human Capital and Health. American journal of human biology : the official journal of the Human Biology Council 2017;29(2) doi: 10.1002/ajhb.22952

3. UNICEF, WHO, World Bank Group. Levels and Trends in Child Malnutrition. In: edition JCME, ed., 2017.

4. Black MM, Walker SP, Fernald LCH, et al. Early childhood development coming of age: science through the life course. Lancet (London, England) 2017;389(10064):77–90. doi: 10.1016/S0140-6736(16)31389-7

5. Das JK, Salam RA, Hadi YB, et al. Preventive lipid-based nutrient supplements given with complementary foods to infants and young children 6 to 23 months of age for health, nutrition, and developmental outcomes. Cochrane Database of Systematic Reviews 2019(5) doi: 10.1002/14651858.CD012611.pub3

6. Prado EL, Larson LM, Cox K, et al. Do effects of early life interventions on linear growth correspond to effects on neurobehavioural development? A systematic review and meta-analysis. The Lancet Global Health 2019;7(10):e1398–e413. doi: 10.1016/S2214-109X(19)30361-4

7. Humphrey JH, Jones AD, Manges A, et al. The Sanitation Hygiene Infant Nutrition Efficacy (SHINE) Trial: Rationale, Design, and Methods. Clin Infect Dis 2015;61 Suppl 7:S685–702. doi: 10.1093/cid/civ844 [published Online First: 2015/11/26]

8. Humphrey JH, Mbuya MNN, Ntozini R, et al. Independent and combined effects of improved water, sanitation, and hygiene, and improved complementary feeding, on child stunting and anaemia in rural Zimbabwe: a cluster-randomised trial. The Lancet Global Health 2019;7(1):e132–e47. doi: 10.1016/S2214-109X(18)30374-7

9. Gladstone MJ, Chandna J, Kandawasvika G, et al. Independent and combined effects of improved water, sanitation, and hygiene (WASH) and improved complementary feeding on early neurodevelopment among children born to HIV-negative mothers in rural Zimbabwe: Substudy of a cluster-randomized trial. PLOS Medicine 2019;16(3):e1002766. doi: 10.1371/journal.pmed.1002766

10. Prendergast AJ, Chasekwa B, Evans C, et al. Independent and combined effects of improved water, sanitation, and hygiene, and improved complementary feeding, on stunting and anaemia among HIV-exposed children in rural Zimbabwe: a cluster-randomised controlled trial. The Lancet Child & Adolescent Health 2019;3(2):77–90. doi: 10.1016/S2352-4642(18)30340-7

11. Chandna J, Ntozini R, Evans C, et al. Effects of improved complementary feeding and improved water, sanitation and hygiene on early child development among HIV-exposed children: substudy of a cluster randomised trial in rural Zimbabwe. BMJ Glob Health 2020;5(1):e001718. doi: 10.1136/bmjgh-2019-001718 [published Online First: 2020/03/07]

12. Piper JD, Mazhanga C, Mapako G, et al. Characterising school-age health and function in rural Zimbabwe using the SAHARAN toolbox. medRxiv 2021:2021.09.22.21263533. doi: 10.1101/2021.09.22.21263533

13. Moss C, Kuche D, Bekele TH, et al. Precision of Measurements Performed by a Cadre of Anthropometrists Trained for a Large Household Nutrition Survey in Ethiopia. Curr Dev Nutr 2020;4(9):zaa139. doi: 10.1093/cdn/nzaa139 [published Online First: 2020/09/15]

14. Ocansey ME, Adu-Afarwuah S, Kumordzie SM, et al. Prenatal and postnatal lipid-based nutrient supplementation and cognitive, social-emotional, and motor function in preschool-aged children in Ghana: a follow-up of a randomized controlled trial. The American journal of clinical nutrition 2019;109(2):322–34. doi: 10.1093/ajcn/nqy303 [published Online First: 2019/02/06]

15. Campbell MK, Piaggio G, Elbourne DR, et al. Consort 2010 statement: extension to cluster randomised trials. BMJ : British Medical Journal 2012;345:e5661. doi: 10.1136/bmj.e5661

16. Clare PJ, Dobbins TA, Mattick RP. Causal models adjusting for time-varying confounding-a systematic review of the literature. International journal of epidemiology 2019;48(1):254–65. doi: 10.1093/ije/dyy218 [published Online First: 2018/10/26]

17. Prendergast AJ, Szubert AJ, Berejena C, et al. Baseline Inflammatory Biomarkers Identify Subgroups of HIV-Infected African Children With Differing Responses to Antiretroviral Therapy. J Infect Dis 2016;214(2):226–36. doi: 10.1093/infdis/jiw148 [published Online First: 2016/05/18]

18. Mundfrom DJ, Shaw DG, Ke TL. Minimum Sample Size Recommendations for Conducting Factor Analyses. International Journal of Testing 2005;5(2):159–68. doi: 10.1207/s15327574ijt0502_4

19. MacCallum RC, Widaman KF, Zhang S, et al. Sample size in factor analysis. Psychological Methods 1999;4(1):84–99. doi: 10.1037/1082-989X.4.1.84

20. Prado EL, Abbeddou S, Adu-Afarwuah S, et al. Predictors and pathways of language and motor development in four prospective cohorts of young children in Ghana, Malawi, and Burkina Faso. Journal of Child Psychology and Psychiatry 2017;58(11):1264–75. doi: 10.1111/jcpp.12751

21. Masquelier B, Hug L, Sharrow D, et al. Global, regional, and national mortality trends in older children and young adolescents from 1990 to 2016: an analysis of empirical data. The Lancet Global Health 2018;6(10):e1087–e99. doi: 10.1016/S2214-109X(18)30353-X

22. Prentice AM, Ward KA, Goldberg GR, et al. Critical windows for nutritional interventions against stunting. The American journal of clinical nutrition 2013;97(5):911–18. doi: 10.3945/ajcn.112.052332

23. Rodriguez-Martinez A, Zhou B, Sophiea MK, et al. Height and body-mass index trajectories of school-aged children and adolescents from 1985 to 2019 in 200 countries and territories: a pooled analysis of 2181 population-based studies with 65 million participants. The Lancet 2020;396(10261):1511–24. doi: 10.1016/S0140-6736(20)31859-6

24. Lelijveld N, Seal A, Wells JC, et al. Chronic disease outcomes after severe acute malnutrition in Malawian children (ChroSAM): a cohort study. The Lancet Global health 2016;4(9):e654–62. doi: 10.1016/s2214-109x(16)30133-4 [published Online First: 2016/07/30]

25. Rosala-Hallas A, Bartlett JW, Filteau S. Growth of HIV-exposed uninfected, compared with HIV-unexposed, Zambian children: a longitudinal analysis from infancy to school age. BMC Pediatrics 2017;17(1):80. doi: 10.1186/s12887-017-0828-6

26. Boivin MJ, Barlow-Mosha L, Chernoff MC, et al. Neuropsychological performance in African children with HIV enrolled in a multisite antiretroviral clinical trial. AIDS (London, England) 2018;32(2):189–204. doi: 10.1097/QAD.0000000000001683

27. Kerr SJ, Puthanakit T, Vibol U, et al. Neurodevelopmental outcomes in HIV-exposed-uninfected children versus those not exposed to HIV. AIDS Care 2014;26(11):1327–35. doi: 10.1080/09540121.2014.920949

